# Longitudinal tracking of inflammatory cell infiltration in the retinal ganglion cell layer in multiple sclerosis patients using high-resolution imaging

**DOI:** 10.1101/2025.04.16.25325926

**Authors:** Elena Gofas-Salas, Mathieu Mossad, Ysoline Beigneux, Nathaniel Norberg, Daniela Castro Farias, Catherine Vignal, Michel Paques, Céline Louapre, Kate Grieve

## Abstract

Neuroinflammation is a critical factor in multiple sclerosis (MS), but *in vivo* monitoring of its cellular dynamics remains a significant challenge. The retina, derived from CNS tissue, provides a unique, non-invasive window into these processes. High-resolution retinal imaging techniques, such as adaptive optics scanning laser ophthalmoscopy (AOSLO), enable the observation of cellular dynamics in the retina, advancing our understanding of neuroinflammation in MS. In this prospective cohort study, we used custom-modified AOSLO to examine the retinal ganglion cell layer in 51 MS patients with different phenotypes: relapsing-remitting MS (RMS) with recent optic neuritis (ON) (N=31), RMS without recent ON (N=12), and progressive MS (N=8), alongside nine healthy controls. We identified immune infiltrates for the first time, likely lymphocytes and microglial cells, near the retinal vascular plexus, with the highest densities observed in ON-RMS patients (93.6%). These cells were sometimes detected weeks before clinical ON onset, and their density declined post-ON, though at varying rates. Infiltrates were more frequently found in MS patients than in controls, even outside acute ON episodes. The cells showed minimal movement and often interacted with vessels, suggesting migratory behavior. Our results suggest that AOSLO imaging can detect subtle retinal inflammatory changes not captured by conventional clinical systems, offering a promising tool for monitoring neuroinflammation in MS and other neurodegenerative diseases. These findings support the potential of high-resolution retinal imaging as a non-invasive biomarker for tracking neuroinflammation and therapeutic responses in patients with MS.

## INTRODUCTION

Neuroinflammation plays a critical role in the progression of multiple sclerosis (MS), a disease characterized by immune-mediated damage to the central nervous system (CNS), leading to demyelination, neuronal loss, and progressive disability. While inflammation contributes to myelin and neuronal damage, it also plays a dual role in promoting tissue repair and regeneration, as evidenced by the activity of microglial cells—the primary innate immune cells of the CNS.^1,2^ Microglia remove cellular debris and aid in recovery following injury, but when this process is disrupted or when microglia maintains a proinflammatory phenotype, regeneration is delayed, emphasizing their dual role in both neuroprotection and neurotoxicity.^3,4^ It is for this reason that a better understanding of how immune responses are implicated in neuronal damage and regeneration is crucially important to understand neurodegenerative diseases and particularly in MS progression.^4^

Recent advances in *in vivo* imaging techniques, such as positron emission tomography and magnetic resonance imaging (MRI), have enabled researchers to track immune activation and microglial responses in the CNS.^5,6^ However, these methods are often limited in their spatial resolution and their ability to capture the full complexity of neuroinflammation, highlighting the need for more precise biomarkers. Identifying specific biomarkers for diffuse inflammation in MS could help to characterize neuroinflammation and monitor disease progression. These biomarkers, potential predictors of disease outcome, could be used to screen patients with a particular phenotype for clinical trials, and could be used eventually as outcome for clinical trials targeting neuroinflammation.^5,6^ The retina, embryonically derived from neural tissue, offers a unique, non-invasive window into CNS pathology, including MS.^7^ The eye has been recognized as a site of injury in MS, with conditions such as retinal phlebitis and damage to the optic nerve being reported.^8^ Like the CNS, the retina contains resident immune cells, primarily microglia, which exhibit dynamic phenotypes ranging from neuroprotective to neurotoxic roles.^9,10^ Clinically, the optic nerve’s involvement in MS is often assessed using optical coherence tomography (OCT), which monitors the damage of the retinal ganglion cells (RGC) using retinal nerve fiber layer (RNFL) and ganglion cell layer (GCL) thickness, as well as macular layers thickness and volume.^11^ While OCT can detect biomarkers like RNFL thinning, its limited resolution means these changes are only identified after significant damage has occurred. Furthermore, OCT images lack the cytological detail needed for accurate identification of cell types, limiting our ability to understand the cellular mechanisms underlying MS-related retinal damage. This highlights the need for high-resolution imaging to capture cellular dynamics earlier in the disease process.

The advancement of imaging technologies in ophthalmology, such as adaptive optics (AO), has begun to bridge the gap in understanding retinal changes associated with these diseases. AO has been integrated into traditional retinal imaging techniques, including OCT, allowing for real-time correction of ocular aberrations and significantly enhancing resolution, which leads to cellular-scale imaging of the retina.^12^ As a result, key cellular changes in retinal tissue have been detected in MS patients using AOOCT, particularly in those with acute optic neuritis (ON).^13^ *In vivo* imaging has revealed RGC degeneration and the presence of macrophage-like cells at the vitreomacular interface, providing valuable insights into the cellular pathology of MS.^13^ While these findings align with pathological studies in the literature,^14,15^ AOOCT technology remains insufficient for detecting all inflammatory cellular features seen in MS histopathology. Histological evaluations by Green et al.^14^ have identified persistent perivascular inflammation and cellular infiltrates in the inner retina, particularly in eyes with a history of ON. Interestingly, this study also showed that similar, though less pronounced, inflammatory changes have been observed in eyes without a history of ON, suggesting a continuous inflammatory process despite the absence of myelin in retinal tissue. Although some putative immune cells have been detected at the border of the RNFL with the vitreous, these perivascular cells seem to remain undetected in AOOCT images.^13^ A significant challenge in retinal imaging is that many cells are either transparent or exhibit low contrast, complicating visualization with conventional ophthalmic imaging modalities, including high-resolution techniques like AOOCT. Adaptive optics scanning laser ophthalmoscopy (AOSLO),^16,17^ a high-resolution imaging technique similar to AOOCT, offers a potential solution. In addition to using AO technology for cellular resolution, the AOSLO has been optimized with an off-axis detection to improve sensitivity to fine phase variations of light passing through transparent or low-contrast cells.^18–22^ This advancement has enabled *in vivo* imaging of RGC and microglial cells in the inner retina in humans in previous studies,^21,22^ making AOSLO a promising tool for the cellular characterization of MS inflammation related retinal changes.

The goal of this study is to identify novel retinal cellular biomarkers of inflammation in MS, with a specific focus on whether retinal inflammation is detectable beyond acute optic neuritis episodes. We used AOSLO phase contrast imaging to monitor the GCL in patients with MS (pwMS), observing for the first time heterogeneous immune cells in the inner retina in MS, particularly after an acute ON. By correlating these findings with clinical symptoms and other imaging modalities like OCT, we aimed to characterize retinal inflammation at different MS stages.

## RESULTS

### Clinical records

Detailed flow chart of inclusion and data analysis is presented in **Supplementary Fig. 1**. Among the total number of participants included in ON-STIM and RETIMUS protocols, 73 subjects were examined with the AOSLO, and from those imaged, 60 participants had images of good enough quality in enough regions of the retina to be included in the study analysis (51 pwMS and 9 healthy controls (HC)). Reasons for poor quality data included poor fixation or high refractive correction. MS patients were categorized in this study into three groups: RMS patients with recent optic neuritis in the last 6 months (ON-RMS, N=31), RMS patients without recent optic neuritis (NON-RMS, N=12); Progressive MS patients without recent optic neuritis (PMS, N=8). The demographic and clinical characteristics of subjects are summarized in **Table 1**. Of these 60 subjects, 40 were women (66.7%), and the mean age was 38.6 years (SD = 11.2). For ON-RMS, 31 affected eyes and 24 fellow eyes were analyzed. In the NON-RMS group, 21 eyes were analyzed, and 15 eyes were analyzed for PMS. The HC group contributed 16 eyes for analysis (see **Supplementary Fig. 1**). Age differed between groups, ranging from 32.74 years (SD = 8.96) in ON-RMS patients to 51.9 years (SD = 8.6) in the PMS group, consistent with known disease demographics. Past episodes of ON occurring more than six months from inclusion were recorded in 5 patients (8.3%), with 2 in the ON-RMS group and 3 in the NON-RMS group. Regarding treatment, 68.6% of patients were receiving disease-modifying therapies.

**Table 1.**
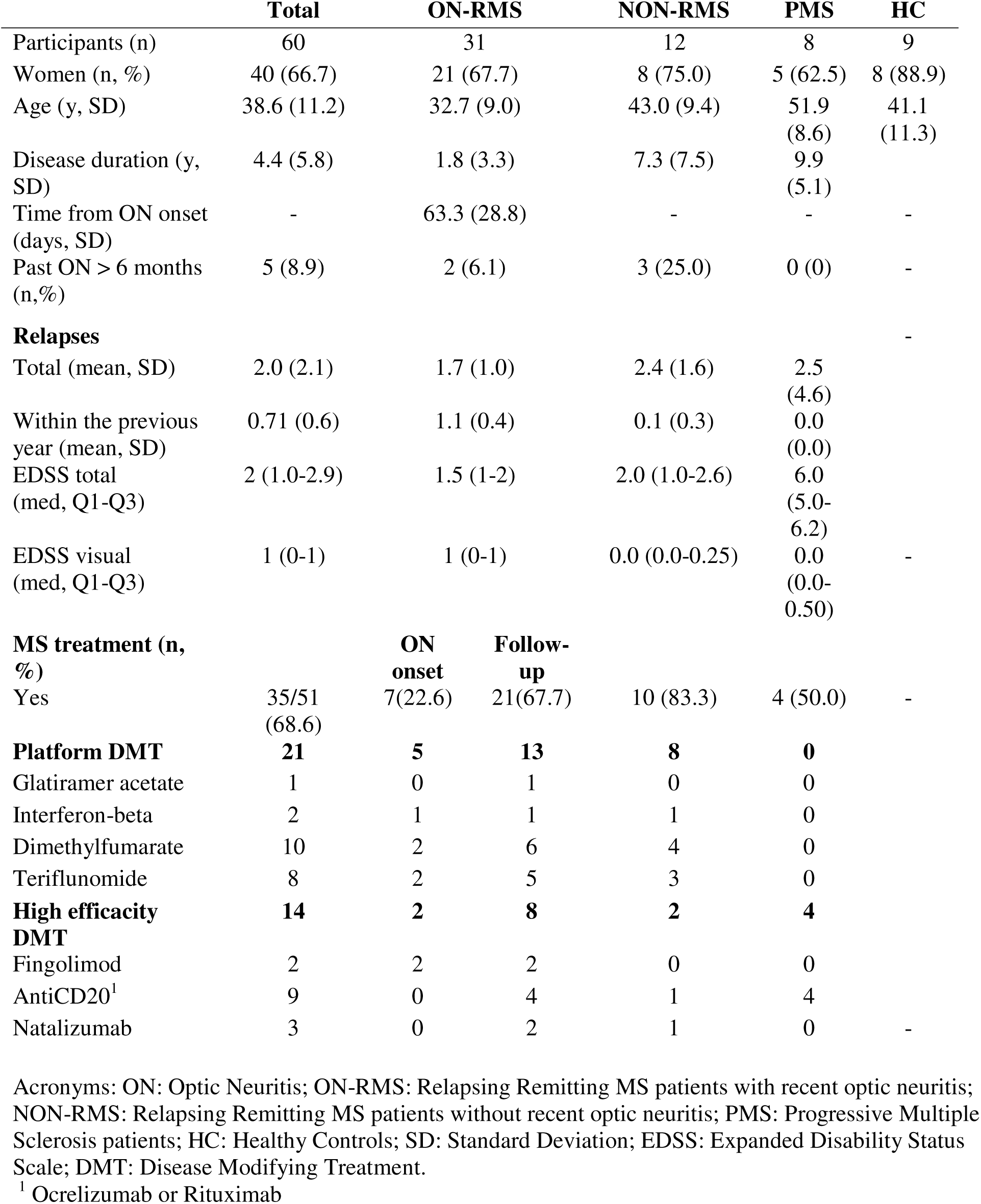
Demographic and clinical data.

The clinical test results of participants are summarized in **Table 2**. Among all groups, worse visual acuity (ETDRS) was observed in the affected eyes of ON-RMS patients compared to their fellow eyes, NON-RMS and HC (*p* < 0.001 for all groups). Similarly, the PMS group showed overall lower visual acuity compared to HC, NON-RMS and ON-RMS fellow eyes (*p* < 0.001 for all groups). Regarding OCT metrics, macular GCL thickness was significantly lower in ON-RMS affected eyes compared to their fellow eyes, NON-RMS, and HC groups (*p* < 0.001). PMS patients also exhibited a significantly thinner GCL layer than HC and ON-RMS fellow eyes (*p* <0.0001). MRI data revealed optic nerve lesions in 81.0% of pwMS. Ninety-seven percent of ON-RMS patients exhibited optic nerve lesions on the side of optic neuritis, compared to only 16.1% of lesions in their fellow eyes’ optic nerve. In contrast, no lesions were found in healthy subjects’ optic nerve.

**Table 2.** Ophthalmological and MRI.

|  | Total | ON-RMS |  | NON-RMS | PMS | HC |
| --- | --- | --- | --- | --- | --- | --- |
|  |  | Affected eye | Fellow eye |  |  |  |
| Visual parameters |  |  |  |  |  |  |
| ETDRS (SD) | 86.1 (9.4) | 79.8 (14.9) | 88.8 (4.9) | 89.7 (4.2) | 84.1 (3.8) | 88.8 (2.9) |
| Sloan 2.5% (SD) | 28.2 (12.6) | 15.6 (13.7) | 34.8 (6.1) | 31.0 (10.2) | 20.5 (11.4) | 35.9 (8.0) |
| VF FT (SD) | 35.0 (5.6) | 30.3 (10.6) | 37.4 (1.6) | 35.5 (3.3) | 34.4 (4.1) | 36.7 (1.2) |
| OCT |  |  |  |  |  |  |
| GCL (SD) | 1.04 (0.19) | 0.89 (0.18) | 1.10 (0.12) | 1.04 (0.18) | 0.96 (0.28) | 1.13 (0.07) |
| pRNFL-m (SD) | 94.26 (16.54) | 88.42 (17.80) | 99.54 (10.45) | 96.96 (20.29) | 85.63 (19.62) | 98.36 (6.92) |
| pRNFL-t (SD) | 63.23 (18.47) | 54.42 (16.60) | 66.18 (13.49) | 65.04 (17.69) | 53.00 (17.13) | 81.33 (17.34) |
| Optic nerve MRI |  |  |  |  |  |  |
| Optic nerve lesion (n,%) | 47/51 (81.0) | 30/31 (96.8) | 5/31 (16.13) | 7/12 (58.3) | 7/8 (87.5) | NA |
| Intraorbital lesion (n, %) | 30/47 (63.8) | 18/30 (60.0) | 3/5 (60.0) | 4/7 (57.1) | 6/7 (85.7) |  |
| Lesion length (mm, SD) | 12.1 (5.5) | 14.2 (4.7) | 6.2 (1.3) | 11.3 (6.6) | 8.9 (4.3) |  |

Acronyms: ON-RMS: Relapsing Multiple Sclerosis (MS) with recent acute Optic Neuritis (ON) ; NON-RMS : Relapsing Multiple Sclerosis without recent acute optic neuritis; PMS : Progressive Multiple Sclerosis; HC : Healthy Controls; VF FT : Visual Field Foveal Threshold; GCL : Ganglion Cell Layer; pRNFL-m/t : peripapillary Retinal Nerve Fiber Layer – mean/temporal

### Cellular infiltrates in the inner retina of MS patients

Cellular infiltrates in the inner retina were detected with AOSLO across all MS subtypes (**Fig. 1**). Many of these cells exhibited lymphocyte-like shapes (**Fig. 2**). In the ON-RMS group, 93.6% of patients presented with cellular infiltrates, compared to 66.7% in NON-RMS and 62.5% in PMS group. **Figure 2** illustrates the morphological characteristics of cells observed in the GCL in most patients, particularly in the affected eye of ON-RMS cases. Most detected cells displayed a round, lymphocyte-like morphology, featuring compact cytoplasm and a smaller, round structure resembling a nucleus (**Fig. 2 F3**). In some instances, only a single intracellular structure was visible, while in others multiple intracellular features were observed, varying in size and position within the cell (**Fig. 2 H1-H5**). These features could represent nuclei or phagocytosed debris and were distinguishable by our system off axis detection due to differences in tissue composition and refractive index. Additionally, **Fig. 3** displays cells with diverse morphologies from both RMS and PMS subtypes, along with HC. Some of these cells are smaller and do not have the compact, round cytoplasm, and their intracellular features are not visible. We detected a low number of round and ovoid small cells in three healthy controls (33.3%), similar to some observed in ON-RMS and NON-RMS patients. Larger cells with one or more round intracellular structures resembling lymphocytes were not observed in healthy controls (**Fig. 3**). Cell diameters ranged from 5.8 µm to 35.6 µm, with a mean diameter of 12.0 ± 4.2 µm across all subjects. Despite size variations, a large portion of cells maintained consistent overall morphology (**Fig. 2 F1-F3**). Further details on the distribution and morphological characteristics of these immune cells are presented in **Fig. 2 (E1-E2)** and **Supplementary Table 1**.

**Fig. 1.**
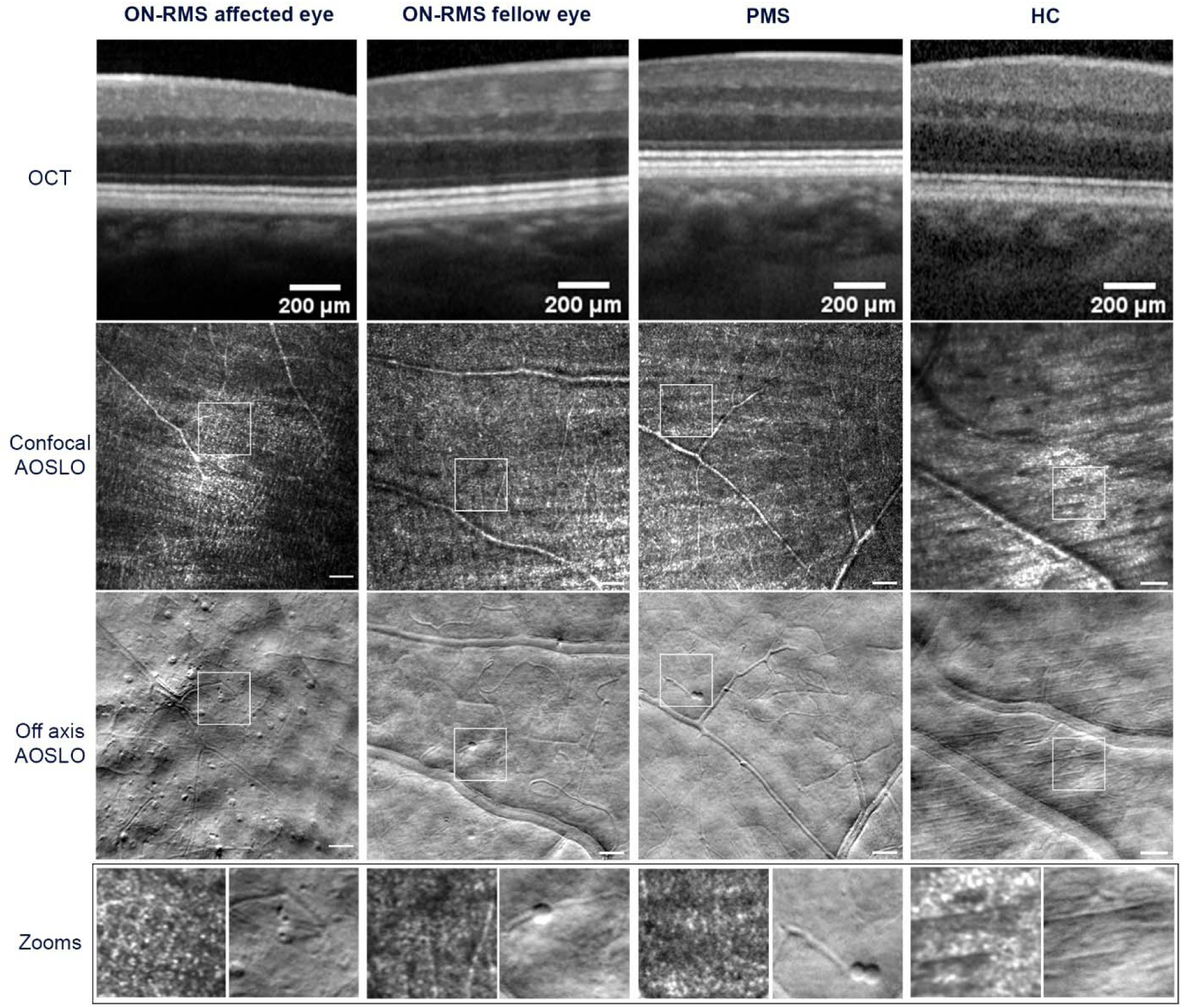
Clinical images of the inner retina from patients with multiple sclerosis (MS) and a healthy control (HC). From the top row to bottom: (1) 4° field of view cross-section of Spectralis OCT images at 4° nasal +/-2°; (2) Confocal and (3) Off-Axis Adaptive Optics Scanning Laser Ophthalmoscope (AOSLO) with a 2° x 2° field of view at 4 ° nasal+/-2°; (4) Zoomed-in images from the confocal and off-axis AOSLO show nerve fibers, capillaries, and cellular infiltrates with lymphocyte-like shape. Scale bars represent 50 µm. From left to right, images from four different subjects: ON-RMS affected eye (Relapsing-Remitting MS with recent optic neuritis), left eye; ON-RMS fellow eye (Relapsing-Remitting MS fellow eye to optic neuritis), right eye; PMS (Progressive MS), left eye; Healthy Control, right eye.

**Fig. 2.**
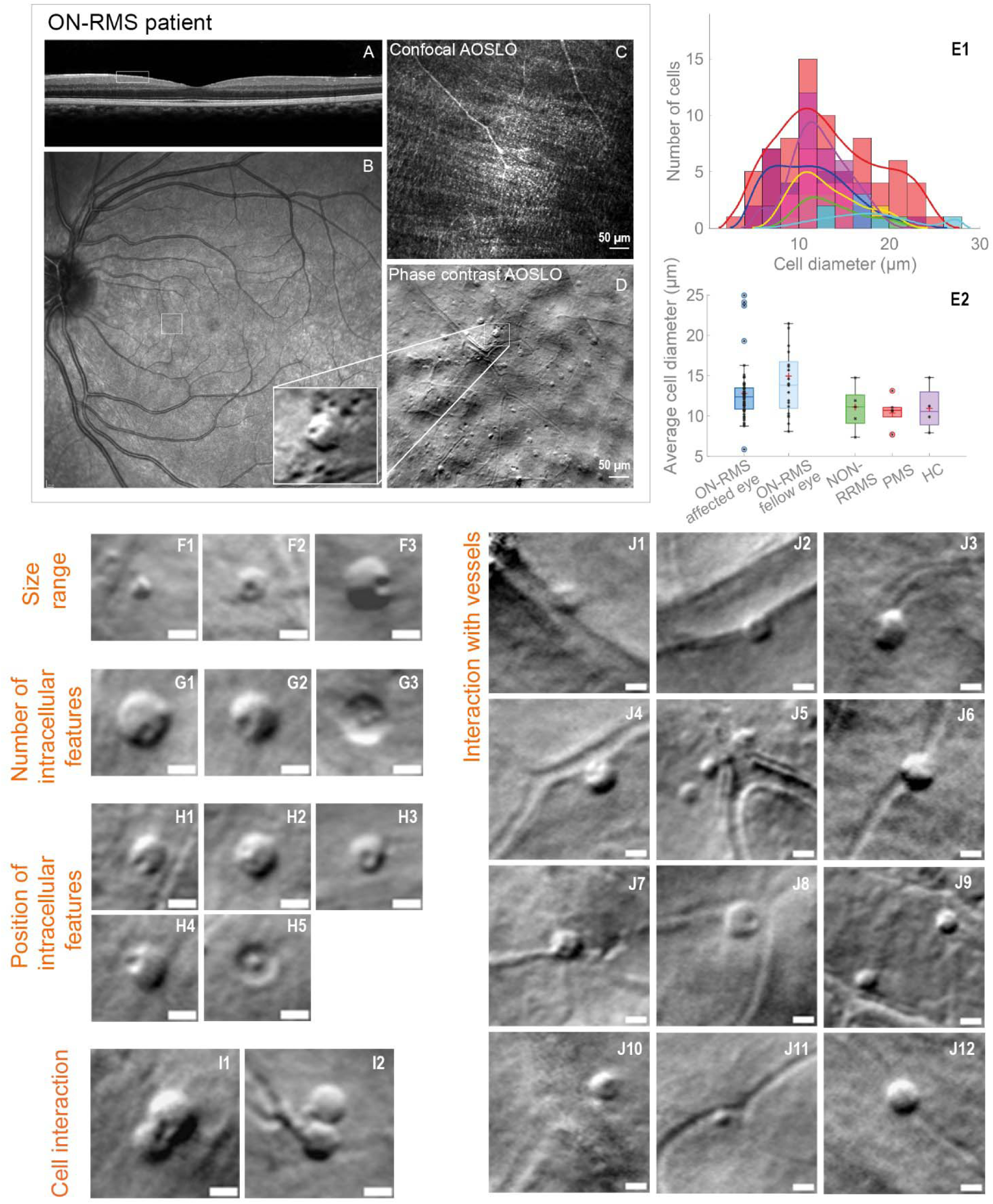
Morphological characterization of cellular infiltrates in the affected eye of ON-RMS patients. (A) OCT (Spectralis) cross-section showing the imaged area (white rectangle), and (B) Scanning Laser Ophthalmoscope (Spectralis) fundus view. (C) Confocal and (D) Off-Axis Adaptive Optics Scanning Laser Ophthalmoscope (AOSLO) images of an ON-RMS patient affected eye on their first visit. Zoomed-in areas highlight lymphocyte-like cells in the layer. Scale bars are 50 µm. (E1) Histogram showing cell count distribution by cell diameter for five ON-RMS affected eyes and one ON-RMS fellow eye. Histogram fits show the size distribution in these patients. (E2) Distribution of average cell size (diameter) for different MS forms (ON-RMS: Relapse-Remitting MS with recent optic neuritis, NON-RMS: Relapse-Remitting MS without recent optic neuritis, PMS: Progressive MS) and Healthy Control (HC). (F1-F3) Infiltrated cell size range in an ON-RMS patient’s affected eye on first visit, 8 weeks after ON diagnosis. (G1-G3) Intracellular features, possibly nuclei, appearing as either single or multiple structures in different cases (e.g., G2: single, G1-G3: multiple). Images from an ON-RMS patient affected eye, 8 weeks after ON (G1-G2) and 29 weeks after ON (G3). (H1-H5) Nuclei-like features in different positions inside the cell. Images from two ON-RMS patients: 7 weeks (H5) and 8 weeks (H1, H2, H4), after ON in one patient, and 8 weeks (H3) after ON in another patient. (I1, I2) Two cells potentially interacting. Images from ON-RMS patient 8 and 29 weeks after ON. (J1-J12) Cell interactions with retinal vessels, some appearing half in the vessel and half in the tissue, suggesting trans-endothelial migration through the vascular plexus. These cells likely represent active infiltration from the vascular system into the inner retina, indicating possible crossing of the blood-retina barrier. Images from five ON-RMS patients at various visits (J1, J12: 5 weeks after ON; J11, J2: 10 and 13 weeks after ON; J3, J6: 18 weeks after ON; J4, J10: 18 weeks after ON; J5, J7, J8, J9: 8 weeks after ON). Scale bars on panels are 10 µm.

**Fig. 3.**
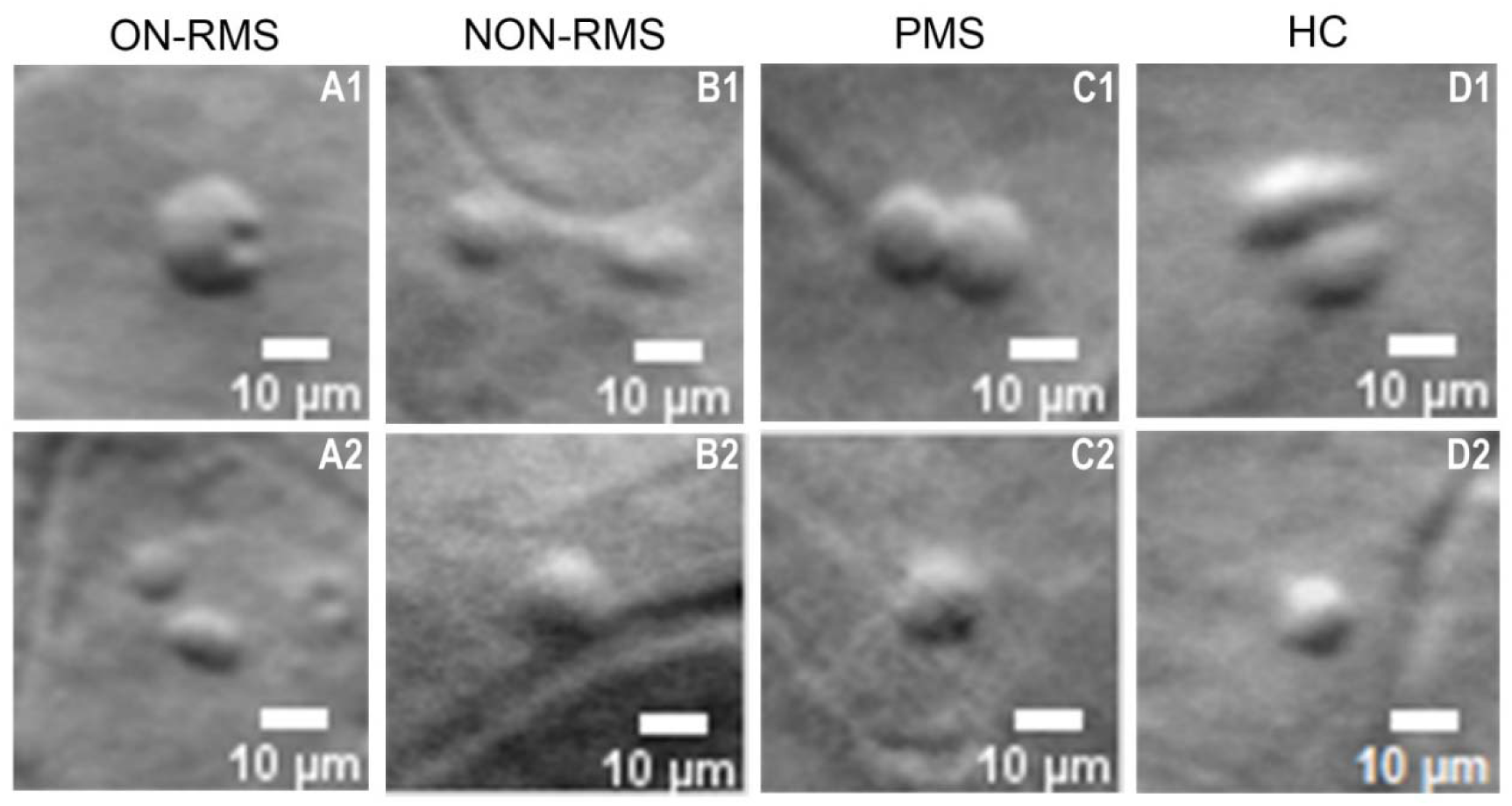
Representative off-axis AOSLO images of putative immune cells with various morphologies in different patient groups and healthy controls: ON-RMS (A1, A2), NON-RMS (B1, B2), PMS (C1, C2), and Healthy Controls (D1, D2). The images show cells resembling innate immune cells (smaller, with less defined and blurrier cytoplasm borders) or lymphocyte-like cells (rounder, larger, with well-defined cell borders, often containing a round intracellular feature), showing morphological variations across groups. Scale bars represent 10 µm.

These cells were predominantly localized in the inner retina, specifically in the GCL, near the RNFL, and just above the vascular plexus. **Supplementary Video 1** demonstrates how these cells become blurred as the focal plane moves axially from the RNFL towards the photoreceptors, just before the vascular structures lose focus. No cells were detected in the outer retina, including the photoreceptor and retinal pigment epithelium layers, across all patients where axial focus shifting was performed. Cells were uniformly detected at various eccentricities, up to 10° from the fovea, in a subset of patients, suggesting a consistent distribution across the retinal eccentricities (see **Supplementary Fig. 2**). We observed a pronounced interaction between the cellular infiltrates and the vascular network, ranging from smaller capillaries to larger vessels. Several examples of these interactions are illustrated in **Fig. 2 J1-J12**. In some instances, the cells appeared to be either partially or fully within the vessels, as if they were entering or exiting the vascular layer. These cells remained stationary for several minutes and, in some cases, up to an hour during the imaging session. A moving cell, highlighted by a red arrow on a vessel, is shown in **Supplementary Video 2**, where its average travel speed was estimated to 3.2+/-1 µm/hour based on this patient data. We noted what appeared to be cellular interactions, as depicted in **Fig. 2 I1-I2**. This phenomenon was captured twice during imaging sessions, each lasting a maximum of 30 minutes. The interactions appeared to be relatively slow, with the cells displaying no noticeable movement over intervals of a few minutes, similarly to their interactions with the vascular structures. Finally, these cellular infiltrates do not produce a confocal signal due to reflection and are therefore not visible in confocal AOSLO images (**Fig 1**). In contrast, several pwMS exhibited hyperreflective dots in the superficial layer of the fovea on confocal images, which lack a phase contrast signal and are absent from phase contrast AOSLO images (see **Supplementary Fig. 3**), confirming that these hyperreflective dots are distinct from the cellular infiltrates described here.

### High cell infiltrate density in acute optic neuritis

We compared the cell density for the different MS groups and for HC. We observed higher cell density in the ON-RMS patients’ affected eye compared to their fellow eyes and compared to all other groups (**Fig. 4A**). Within ON-RMS affected eyes, cell count was substantially heterogeneous, suggesting that inflammatory responses may differ among patients. Moreover, we noticed that ON-RMS patients presented large density variations over time, with a rapid decrease in the weeks following ON (**Fig. 4B**), but the rates of decline varied significantly among individual cases. Of the 31 non-RMS patients, 13 had sufficient data for longitudinal analysis, and for these, a monoexponential decay model was applied. Seven patients demonstrated a valid monoexponential decay model, while the remaining six did not meet the R² threshold. The Kruskal-Wallis test revealed no significant statistical differences, indicating that, overall, the monoexponential decay model adequately describes density dynamics after ON. The half-lives of cell density in patients exhibiting monoexponential decay ranged from 0.52 weeks to 33.5 weeks, with a mean of 8.2 weeks, reflecting considerable variability in decay rates. In some patients, cells were still detected months after the ON episode, but typically at a low density of 3–5 cells per 600 µm² retinal area. Most cellular infiltrates were detected in the affected eyes of ON-RMS patients; however, some cells were also observed in the fellow eyes (**Fig. 1**), while no abnormalities were noted in other cases. The fellow eyes that exhibited cellular presence showed a consistently low cell count, approaching zero but remaining detectable (**Fig. 4 A**). Similarly, a low cell density was observed in NON-RMS and PMS patients, with even lower counts found in healthy controls. Although all ON-RMS patients were imaged following a diagnosis of ON, one patient experienced an acute ON in the fellow eye during the follow-up (**Fig.4 C**). We observed a sudden increase in cell density in the nasal region of this fellow eye 7 weeks before clinical symptoms and before detection of abnormalities in complementary clinical tests, including visual acuity assessments and OCT imaging. This early increase in cell density preceded the subsequent diagnosis of ON. While this is a single case, it suggests that these cells may appear early in the ON episode, prior to the manifestation of clinical symptoms.

**Fig. 4.**
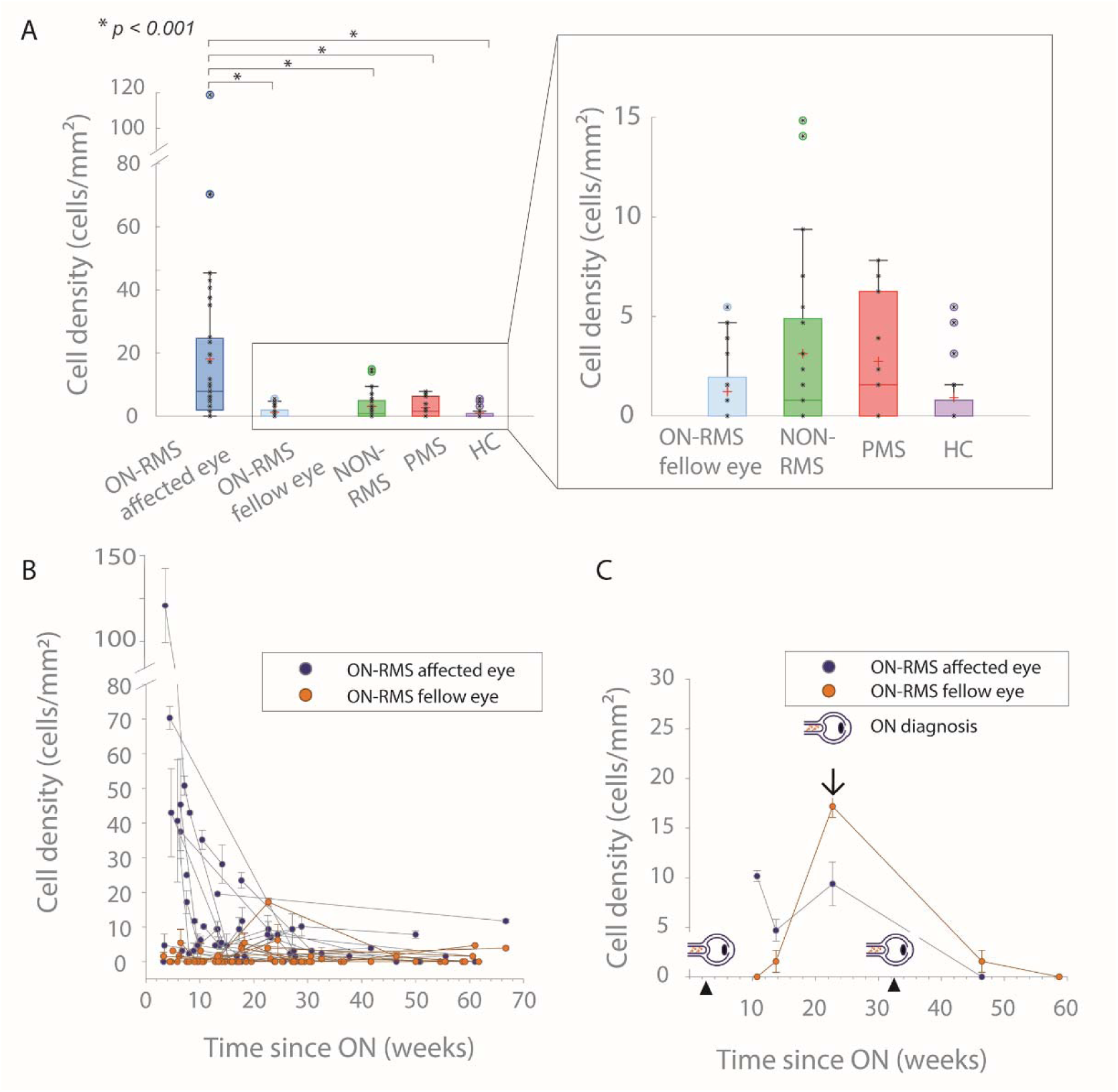
Evaluation of the influence of MS subtype on immune cell density. (A) Cell density at baseline (first visit on the AOSLO) for each MS form (ON-RMS: Optic Neuritis in Relapsing-Remitting MS, NON-RMS: Relapsing-Remitting MS without ON, NON-PMS: Progressive MS without ON) compared to Healthy Controls (HC). One-way ANOVA revealed a significant difference between the affected eye in ON-RMS and all other groups (\**P* < 0.001), but no significant differences between the other groups. (B) Cell density in the affected and fellow eyes of ON-RMS patients over time (in weeks) following ON diagnosis. (C) Cell density in both the affected and fellow eyes of an ON-RMS patient, tracked over several weeks following optic neuritis (ON) diagnosis. The arrow indicates marked increase in density preceding the ON diagnosis in the fellow eye by 7 weeks, suggesting early preclinical alterations.

### Absence of association between optic nerve lesion characteristics and cellular infiltrates

We also explored other factors that could influence cell density, beyond the time factor. Specifically, we assessed the relationship between cell density and optic nerve length, distance between papilla and lesion, and intraocular lesion location, accounting for the time elapsed to the ON event. Our analysis revealed no significant correlation between the topography of optic nerve lesion and cellular density. Specifically, the scatter plot illustrating cell density in relation to ON length (**Fig. 5A**) shows no evidence that variations in ON length are associated with changes in cellular density. Furthermore, we did not observe a higher cell density in case of intraorbital lesions versus other location (canalicular, cisternal, or chiasma) (**Fig. 5B**). Similarly, we did not find a correlation between cell density and distance between papilla and optic nerve lesion (**Fig. 5C**). We found no correlation between cell density measured at nasal eccentricities and that measured at temporal eccentricities (**Supplementary Fig. 4**). Finally, we examined the potential influence visual acuity and disease-modifying treatments (DMTs) administered at the time of patient inclusion and the timing of imaging (**Supplementary Fig. 4** and **5**). In ON-RMS affected eye, we did not find any association between retinal cellular density and visual acuity or MS treatments, even after accounting for the time elapsed to the ON event (see **Supplementary Fig.4** and **5**). Interestingly, in one NON-RMS patient on natalizumab, cellular density was high (10-15 cells/mm^2^).

**Fig. 5.**
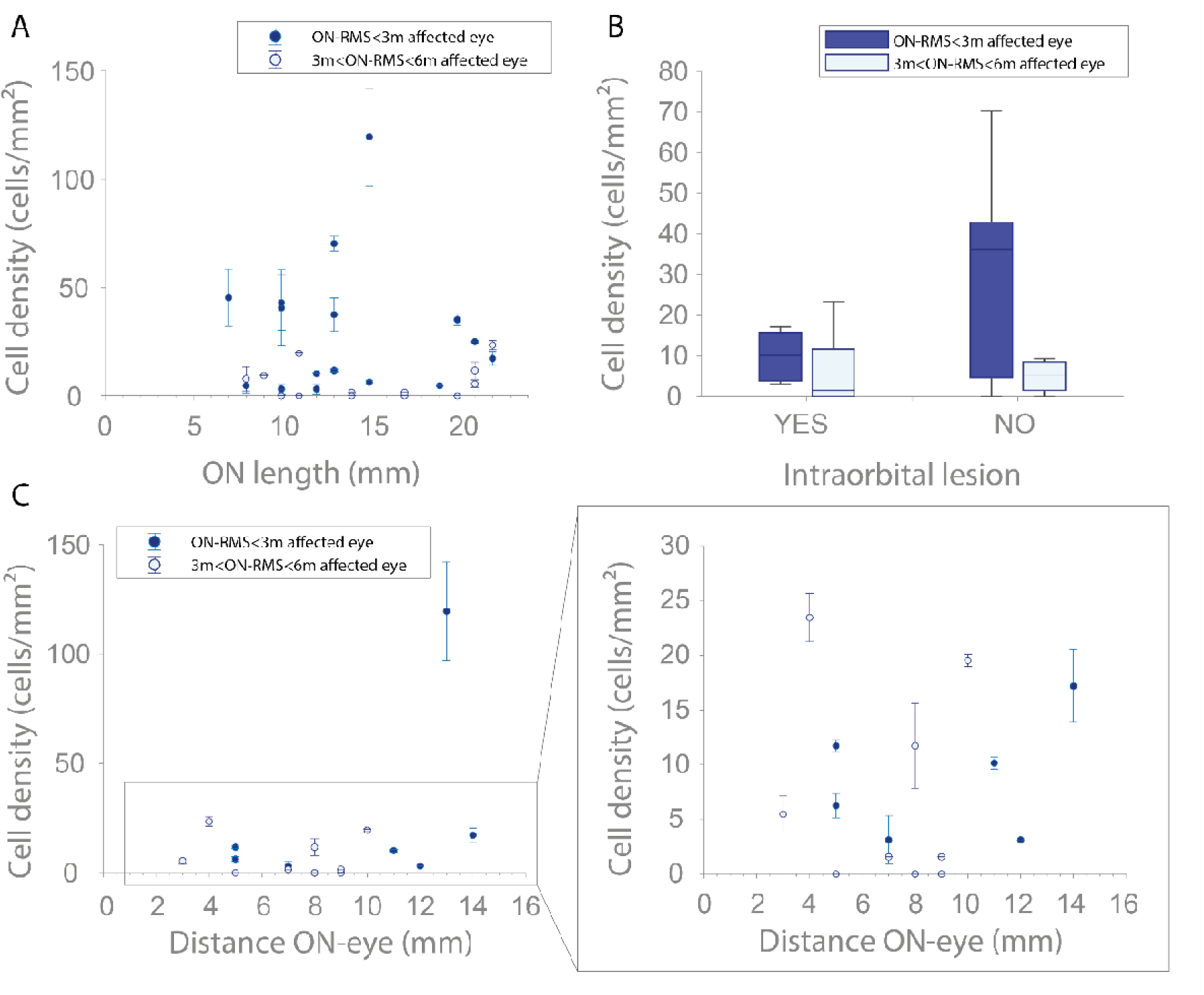
Evaluation of the influence of MRI lesion topography on immune cell density. (A-B) Influence of ON length and intraorbital lesion presence on cell density in the affected eye of ON-RMS patients, assessed at 3 months and 3-6 months post-ON. (C) Effect of ON lesion distance on cell density in ON-RMS patients with an intraorbital lesion (YES category in (B)), evaluated at the same time points. Statistical analysis revealed no significant correlation between cell density and ON length, ON distance, or intra-orbital lesions. No significant effects were found for other factors after adjusting for time to the ON event.

### Cases of highly degenerated retina

In three cases involving pwMS, we observed the presence of degenerative cysts (**Fig. 6**). These cysts sometimes exhibited a circular shape that could be mistaken for lymphocyte-like cells, particularly when they are small. However, we noticed that cysts often have irregular shapes, and their borders appear less compact than those of the cells. Additionally, cyst diameter increases, rapidly exceeding the maximum size observed for the lymphocyte-like cells. **Figure 6** illustrates the progression of cyst formation in a region of the retina over the course of one year. In Fig. 6D, we show the borders of the cysts across three visits, highlighting how their shape continues to change even after they stop growing in size. In areas completely degenerated by cysts, we did not observe any cells; this may be attributed to the fact that cysts likely alter the refractive index of the surrounding tissue, making it difficult to detect cells in that region. Consequently, although cell density was determined to be zero in these cases, it is possible that some cells were present but undetected by the off-axis AOSLO system.

**Fig. 6.**
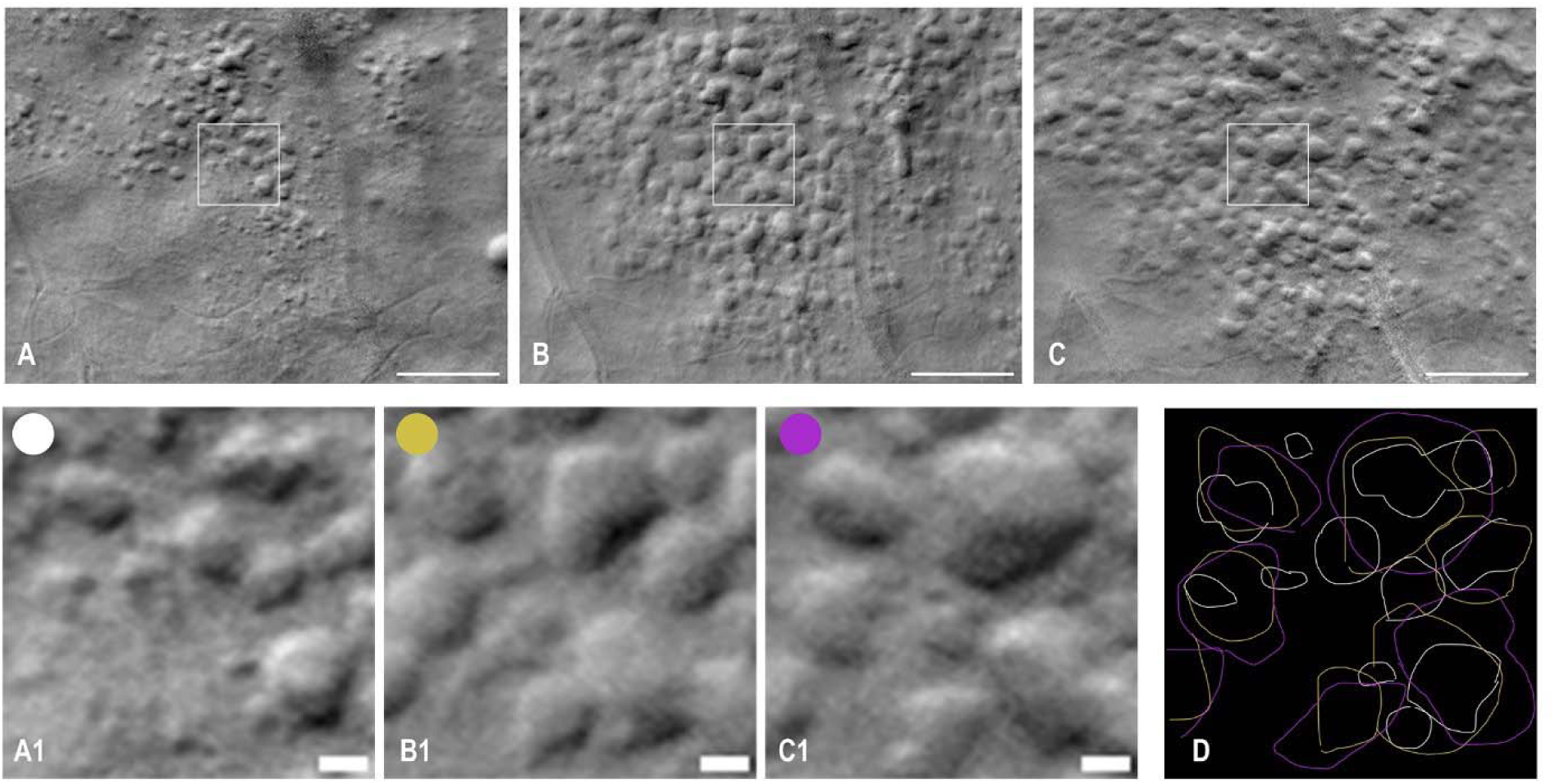
Off-axis Adaptive Optics Scanning Laser Ophthalmoscope (AOSLO) images of degenerative cysts in ON-RMS patient (OS eye), tracked over one year. The images correspond to visits 1, 3, and 4 at 13, 36, and 61 weeks post-ON, respectively. (A1-C1) Magnified regions from (A-C) illustrate the progression of cysts, with some small cysts enlarging while others maintain similar sizes but change in morphology. (D) Overlay of cyst borders from each visit (A1-A2-A3: white, yellow, and purple lines), manually delineated, highlighting the variation in cyst size and shape over time. Scale bars represent (A-C)100µm and (A1-C1) 10 µm respectively.

## DISCUSSION

Using high-resolution adaptive optics retinal imaging in patients with MS, we were able to reveal potential immune infiltrates in the GCL of the retina from the retinal vascular plexus. Morphological features of these cellular infiltrates suggest that they are composed of lymphocytes and microglial cells, in close proximity to vascular structures. The highest density of infiltrates was detected in the retina of patients who experienced a recent ON, and the presence of cellular infiltrate may even precede clinical onset. Cellular density decreases in the months following ON, although the rate of decrease is heterogeneous across patients. Outside acute ON, retinal cellular infiltrates are detectable in pwMS more frequently compared to healthy controls.

The morphology of the cells that we observed in the inner retina aligns with histological findings from the largest retinal pathology study in MS.^14^ This histology series revealed perivascular infiltrates through immunostaining, with most cells displaying round, mononuclear shapes characteristic of lymphocyte cells. However, the authors also observed cells with diverse morphologies, indicating the presence of different immune cell types, similar to our findings. Morphologically, activated B lymphocytes develop into plasma cells and assume a distinctive profile characterized by an eccentric nucleus and a prominent, also eccentrically placed, perinuclear pale zone which could account for the characteristic shape we noticed in these round cells which also have an eccentric intracellular feature. Some of these cells present more than one intracellular grain, which could be multiple nuclei or simply additional intracellular structures like debris. However, some immune cells can present, when active, as granular cells distinguished by their larger overall size with an increased quantity of cytoplasm and various prominent cytoplasmic granules, such as neutrophils.^23,24^ Moreover, using the same AOSLO imaging modality in mice with induced retinal inflammation, Joseph et al.^25^ observed neutrophils rolling along the venular endothelium, as well as infiltrates of monocytes and macrophages both within vessels and extravasated into retinal tissue. These immune cells detected in the ganglion cell layer exhibit a morphology similar to the cellular infiltrates observed in this study. In addition to the similar morphology, the diameters of the cell infiltrates observed in pwMS of our study fall within the size ranges of immune cells, such as lymphocytes (8-15 µm^23^), monocytes (12-20µm^24^), or macrophages (10-30 µm^24^), as reported in the literature. Consistent with previous pathological studies,^14^ these infiltrates were detected solely in the GCL and RNFL, with none observed in the outer retina, and were present at various eccentricities across the retina. Moreover, our phase contrast detection allows for intracellular analysis as it reveals both the outer and inner boundaries of cells due to variations in refractive index. Several of the cells exhibit boundaries resembling those of lymphocytes imaged with refractive index tomography (RIT), a technique that visualizes and quantifies refractive index distribution, providing information about the internal cellular structure.^26^ For example, cells with small, round intracellular features show a refractive index distribution similar to that of B lymphocytes. In contrast, cells lacking visible intracellular structures resemble the more uniform distribution seen in T lymphocytes, as reported in Yoon et al.^26^

In addition to being anatomically alike in shape and distribution, these cells seem to display a similar behavior to immune infiltrates. In several instances, cells appear to enter or exit the medium or even roll inside vessels, consistent with the mechanisms of immune responses in local retinal inflammation.^14^ The dynamic of these cells is slow, going from what seems an immobile state to a few microns displacement per hour. Contrary to animal models where cellular dynamics are more accessible, the motility and speed of retinal immune cells in the human retina has scarcely been studied, and therefore it remains difficult to assess the resemblance in dynamics in animals with our results. However, inflammation progression in humans has been described as very slow (a few microns per month), and depending on their role, some cells can remain in quite fixed positions. In addition, the speed of a few microns per hour estimated from the cell motion in a vessel is in accordance with retinal immune cell velocity from the literature^22^. Another observation suggestive of lymphocyte and macrophage presence is the apparent cell interactions depicted in **Fig. 2 I1-I2**. The cells in contact exhibit characteristics similar to those seen during antigen-presenting cell (APC) signaling to T cells.^27^ Particularly, in **Fig. 2 I1**, one cell appears larger than the other, a change often associated with cellular activation. However, while the intracellular distribution observed in these cells corresponds with those reported for lymphocytes in the literature, other immune cells may exhibit similar refractive index characteristics, complicating definitive identification.

We also observed that while not all MS patients exhibited these cells, 93.6% of ON-RMS patients did, particularly following or around an acute ON episode. These patients displayed various cell morphologies, including numerous lymphocyte-like cells with a rounder, larger structure, well-defined borders, and often a distinct intracellular feature. In contrast, such cells were not observed in the three healthy controls where other cells were detected. Thus, we may be simultaneously detecting cellular infiltrates and innate immune cells already present in the tissue, potentially in their activated form. Previous AOSLO studies have also detected microglia in healthy individuals,^22^ within the RGC layer, which also displayed smooth, round, or elliptical shapes at the smaller end of the size range reported in our study. This may account for the presence of cells in healthy controls and be consistent with the retinal property of immune privilege, which implies the exclusion of leukocytes, including granulocytes, myeloid-derived cells, and lymphocytes, from the tissue.

The similarities with Green et al.^14^ indicate that we were able to detect similar localized inflammatory changes to those observed in their MS samples, but directly in patients using off-axis AOSLO, suggesting we might be observing the same group of cells. Our *in vivo* imaging of these cells in patients addresses some of the significant limitations of traditional histological studies, such as eliminating the issues associated with post-mortem imaging, where physiological changes may alter the observed processes. Additionally, we can image younger patients with varying disease durations and even follow them over time to evaluate the long-term cellular dynamics of this layer.

We observed a significantly higher cell density in eyes recently diagnosed with acute optic neuritis (ON-RMS) compared to other MS patients without recent ON and healthy controls. Hammer et al.^13^ previously reported an enhanced immune response in ON eyes, documenting an increase in macrophage-like cells at the vitreomacular interface, although their observations did not specifically concern the same cell types in the same retinal layer. In our study, the correlation between cell density and ON episodes was further supported in the case of one patient who experienced a sudden increase in cell count in the fellow eye, shortly before being diagnosed with acute ON. This increase occurred prior to typical clinical signs of ON, although the patient did show a slight decrease in low-contrast vision. Thus, these cells may appear before significant signs in other clinical tests, potentially alerting to the risk of ON in MS. Interestingly, in the affected eye of ON-RMS, the same lymphocyte-like cells were observed both early post-ON and months after the episode. The long-term retention of these cells in the retina may suggest a persistent underlying immune response. Similarly, these cells, typically absent from immunoprivileged tissue, were also detected in patients without recent ON. This inference raises the question of how this tissue, which is devoid of myelin, could maintain what seems to be a persistent inflammatory response. Specific microvascular permeability may explain immune infiltration, common to the brain and the retina.

On the other hand, cell densities in the fellow eyes of ON-RMS patients, as well as in NON-RMS and NON-PMS patients, were very low, with many individuals showing no cells at all. The presence of some cells in healthy controls raises questions about establishing a pathological threshold. This observation suggests that, with the current technique, we cannot rely solely on cell density metrics as biomarkers for inflammation unless the cell count is exceedingly high, as seen in ON-affected eyes, or if the cells exhibit significantly different morphology from innate microglia, such as the lymphocyte-like cells. Further research is necessary to characterize these cells, potentially through morphological analysis and machine learning of features. Future studies will focus on this characterization, particularly in comparison with histopathological analyses and animal models. The retina remains underexplored in histology concerning MS, and histological imaging techniques differ significantly in contrast from our clinical system. Subsequent efforts will aim to image our samples using comparable contrast and to label various immune cell types for a deeper exploration of the data from this study.

Regarding disease-modifying treatments (DMTs), while definitive conclusions are difficult to draw, there does not appear to be a significant effect of DMTs on cell density. Although the limited data prevent firm conclusions, an interesting observation was made in one case of a NON-RMS patient treated with natalizumab, where, despite having treatment targeting alpha-4 integrin and therefore limiting leucocyte trafficking through endothelium, a high cell density (greater than 10 cells/mm²) was observed. This questions the pharmacodynamics of such treatments on specific tissue compartments like the retina.

Another challenge in analyzing these cellular infiltrates is the presence of degenerative cysts, especially when the RNFL is severely atrophied. In our observations, cysts, captured approximately six months apart, progressively grow and invade the GCL across various eccentricities. Small cysts may be mistaken for cells, although they can often be distinguished by their irregular, slightly blurry borders. However, as they expand, it becomes possible that cells might be present inside the cysts, but due to changes in the refractive index, they remain undetectable. Despite this, the cysts themselves are clearly visible, appear to evolve over time, even when significantly enlarged and therefore could be interesting to monitor during MS course. Finally, previous studies^28,29^ have reported a presence of hyperreflective foci in various retinal layers using OCT, including the GCL, in pwMS. In our data, confocal AOSLO images, which typically reveal features also visible in OCT (such as cysts or deposits), did not show hyperreflective dots either in these layers, or at the locations of the immune cells described here. Although we observed deposit-like dots exclusively in the fovea of some patients, a finding consistent with previous studies^30^ using confocal AOSLO, these deposits were only present in the fovea and appeared in areas devoid of cells at the same locations. Moreover, the cells observed in our study have a mean size of 12 µm, which is below the standard OCT resolution and thus potentially undetectable in OCT cross-sections. However, expanding this study to investigate a potential correlation between these findings and the hyperreflective foci recently identified in pwMS retina on OCT would be valuable.

In conclusion, our study highlights the potential of phase-contrast AOSLO to provide real-time, *in vivo* visualization of immune responses in the GCL, specifically in pwMS with recent ON. The detection of unique cellular features and the temporal changes in cell density underscore the capability of this technology to monitor inflammatory mechanisms non-invasively. This approach offers a promising avenue for addressing the limitations of conventional histopathology by enabling dynamic monitoring of inflammation associated with MS. Furthermore, these findings suggest that phase-contrast AOSLO could serve as a valuable biomarker for assessing inflammation in MS and potentially other neuroinflammatory conditions. Future research integrating imaging data with systemic inflammatory markers and exploring the functional impact of these immune cells will help advance our understanding of their role in MS progression and therapeutic responses.

## MATERIALS AND METHODS

### Case selection, consent and ethical approval

All subjects in this study were recruited at the Pitié-Salpêtrière Hospital, Paris, and included in two clinical studies: 39 pwMS were included in the ON-STIM Study (ClinicalTrials.gov-ID:NCT04042363) where recruitment of subjects is made at an early stage of an acute ON (within 3 months from ON treatment); 35 pwMS were included in the RETIMUS study (ClinicalTrials.gov-ID:NCT04289909) where they were recruited based on MS clinical course and divided in 3 groups: (1) patients with relapsing remitting MS (RMS) who did not have an acute ON in the past 6 months, (2) patients with RMS who had an acute ON within the last 6 months, (3) patients with primary or secondary progressive MS (PMS).

Healthy controls were included in these protocols, and 31 were recruited during the time of the study, of which 9 were available for AOSLO imaging. Inclusion criteria for healthy controls included age 18–60 years and no diagnosis of ocular pathology.

For the analysis, patients included in RETIMUS study and with ON within 6 months from inclusion were pooled with patients included in ON-STIM study, forming the group of RMS patients with recent ON (ON-RMS group). Depending on the study, we proposed longitudinal imaging with the AOSLO system at 3, 6, and 12 months for the ON-STIM study and at 3 and 6 months for the RETIMUS study. Follow-up AOSLO was performed based on a machine and technician availability. In certain cases, patients with high cell density had additional visits scheduled to capture supplementary acquisitions. AOSLO examinations were carried out between January 2020 and January 2024.

The study was approved by an ethic committee (Comité de Protection des Personnes, CPP) and written informed consent was obtained prior to inclusion. All retinal imaging was performed at light levels conform to ISO and ANSI standards, with near infrared light which is most comfortable for the patient.

### Ophthalmic, neurological, OCT and MRI examinations

Ophthalmic and neurological examinations were performed at each visit, with visual outcomes measure using high-contrast visual acuity (ETDRS, measured by the number of correct letters) and low-contrast visual acuity (Sloan, 2.5%). Eye fundus images and OCT cross-sections were captured using the SPECTRALIS system (Heidelberg, Germany) for all patients and controls at each visit. In addition, all subjects underwent at inclusion MRI on a Siemens Prisma 3T machine, which included brain sequences (3D FLAIR, 3D DIR, T1+gadolinium) and orbital sequences (3D DIR on orbits, 2D proton density, T1 DANTE+gadolinium). Optic nerve lesions were manually segmented, and metrics such as lesion location (intra-orbital, canalicular, cisternal, or chiasma), lesion length, and the distance between the papilla and the lesion (for intraorbital lesions) were extracted.

### Adaptive Optics Scanning Laser Ophthalmoscope

Patients underwent consistent imaging using an AOSLO during each visit. The AOSLO system (a custom modified MAORI device, PSI, Andover MA, USA), installed at the Clinical Investigation Center (CIC) of Quinze-Vingts Hospital, has been previously described in detail by Grieve et al.^31^ In summary, this system utilizes adaptive optics correction to enhance Scanning Laser Ophthalmoscope (SLO) images, providing exceptional spatial resolution capable of imaging individual cone photoreceptors. The AOSLO includes a confocal reflectance channel and four off-axis apertures for non-confocal imaging, with optical fibers acting as apertures to relay light to avalanche photodiode (APD) detectors. A 69-actuator deformable mirror (DM69, Alpao, France), operating in a closed loop at 10 Hz with a Shack-Hartmann wavefront sensor, corrects wavefront aberrations. The off-axis detectors generate phase contrast images that reveal transparent features, such as low-contrast capillaries and certain retinal cells. These apertures are positioned in opposite sides of a central confocal aperture, capturing photons that have been multiply scattered by the deeper layers of the retina and refracted by transparent cells, creating variations in intensity based on their refractive indices. By subtracting and normalizing images from two opposite off-axis apertures, we obtain high-contrast images of retinal cellular structures. Confocal images were simultaneously captured alongside the off-axis AOSLO images to visualize highly reflective cellular structures, such as nerve fibers. The image sequences were corrected for eye motion and averaged to enhance the signal-to-noise ratio. Each of the imaging sub-systems has an independent light source: 757 nm (Exalos SLD; 757 nm center, 20 nm full-width at half maximum (FWHM)) for the AOSLO, and 840 nm for the wavefront sensing beacon. Unless contraindicated by the ophthalmologist, participants received eye drops for pupil dilation and cycloplegia (one drop of mydriaticum 0,5%).

Imaging locations were selected at 4° eccentricity on either side of the fovea, with a ±2° range in all directions. Each location is designated using a naming convention that specifies the degree of eccentricity and direction (T – temporal, N – nasal, S – superior, I – inferior). At each location, we acquired ten image sequences with a field of view of 2° x 2°, focusing on the GCL. Extended imaging was performed for a subset of participants at various retinal eccentricities, up to 10° from the fovea in all directions, and at different depths, ranging from the RNFL to the photoreceptor layer. Additional AOSLO images were acquired at the fovea to assess hyperreflective structures potentially associated with MS. Each imaging sequence consisted of 300 frames captured at 24 frames per second. Participants were imaged at various time intervals (from minutes to weeks) to monitor cellular dynamics in the retinal GCL. Short-term and long-term videos were generated by aligning averaged images either within a single visit or across multiple visits (referred to as timelapse). To accurately localize cellular structures in the retina, images were also aligned through various focal depths in consecutive retinal layers (referred to as depthlapse), with each frame representing a distinct retinal depth. The alignment of both timelapse and depthlapse image stacks was achieved using custom cross-correlation algorithms (Matlab,The MathWorks Inc., Natick, Massachusetts).

### Cellular infiltrates analysis

We analyzed AOSLO off-axis images of the GCL, detecting cellular infiltrates around the retinal vessels. Cell counts were manually performed by two independent experienced raters (authors EGS, KG) using custom ImageJ—Fiji 2.0.0 software, and the average count at each location was calculated between the two raters. The mean number of cells per mm² was used to measure cell density. Cell size was determined by marking the cytoplasm, with both raters contributing to the measurements.

### Statistical analysis

Cell density and average cell size within the regions of interest (ROI) around 4° ± 2° temporally (T) and nasally (N) were compared using Matlab Software (Matlab,The MathWorks Inc., Natick, Massachusetts). We performed a Kruskal-Wallis test to compare cell density between the different groups due to the lack of normal distribution and unequal group sizes. A one-way ANOVA was applied to compare the average cell size between the groups, as the data followed a normal distribution. When the ANOVA showed significant results, we conducted a Tukey-Kramer post-hoc test to assess pairwise comparisons between groups. We analyzed cell density over time in ON-RMS patients by selecting subjects with at least 3 follow-up data points and modeling the decay using a monoexponential curve. Statistical comparisons were then conducted based on the half-lives of the decay curves. Finally, in ON-RMS group, we assessed the relationships between cellular infiltrate on one side, and optic nerve lesion length, distance from papilla, lesion location, and visual acuity on the other side, accounting for the time elapsed to the ON event. Non-parametric Spearman correlation coefficients were computed to test the association between the variables, given the absence of normality in some datasets. Partial correlations were used to adjust for the potential confounding effect of the time elapsed to the ON event. A multivariate robust linear regression model was employed to explore the combined influence of these variables on ON-related outcomes while minimizing the impact of outliers. All analyses were performed using Matlab (Matlab,The MathWorks Inc., Natick, Massachusetts), and statistical significance was set at p < 0.05.

Finally, z-tests were computed to compare clinical results between the different groups.

## List of Supplementary Materials

Fig. S1 to S5

Table S1

Movies S1 to S2

## Data Availability

Data and materials availability: Deidentified raw data presented in this study is available upon reasonable request from the corresponding author for privacy reasons.

## Acknowledgments

We thank Amandine Hippolyte, Annouk Szabo, Carole Romand and Celine Devisme for coordinating clinical investigations. We thank FCRIN4MS (French Clinical Research Investigation Netwoks for MS) and FRCRNET networks for assistance in setting up the protocol and following study progress.

## Data and materials availability

Deidentified raw data presented in this study is available upon reasonable request from the corresponding author for privacy reasons.

