## Supplemental Figures and Table for "Longitudinal tracking of inflammatory cell infiltration in the retinal ganglion cell layer in multiple sclerosis patients using high-resolution imaging"

### Supplementary Figures and Tables

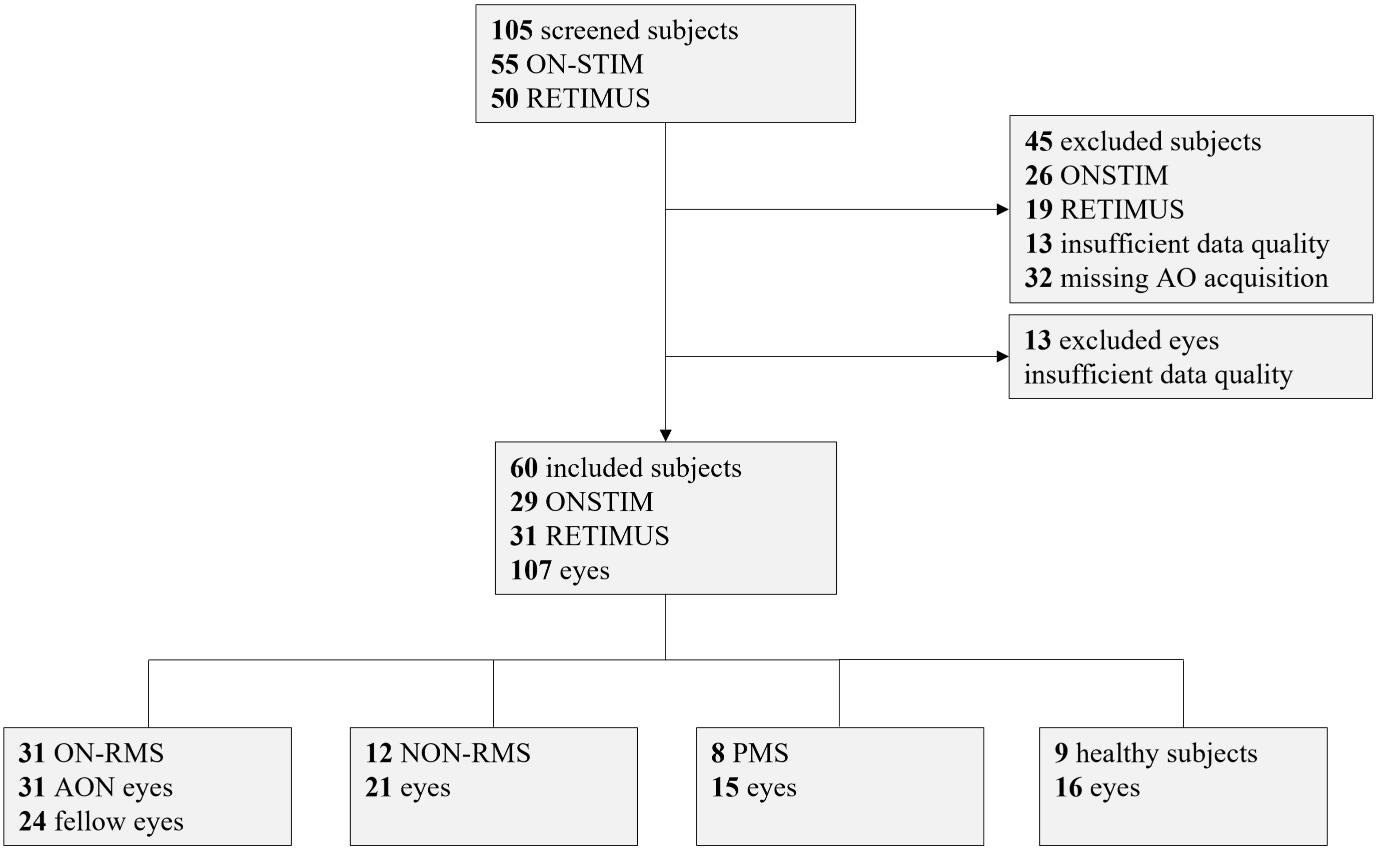


**Supplementary Figure 1.** Flow chart illustrating patient inclusion from two clinical studies, ON-STIM (ClinicalTrials.gov ID: NCT04042363) and RETIMUS (ClinicalTrials.gov ID: NCT04289909), along with reasons for data exclusion. Exclusions include missing AO data due to system upgrades or the absence of a trained imaging technician, as well as poor data quality caused by fixation issues or excessively thick ocular media. The figure also shows the distribution into the four groups of patients with MS and controls. AO: Adaptive Optics; ON-RMS: Relapsing-Remitting Multiple Sclerosis with recent acute optic neuritis; NON-RMS: Relapsing Multiple Sclerosis without recent acute optic neuritis; PMS: Progressive Multiple Sclerosis.

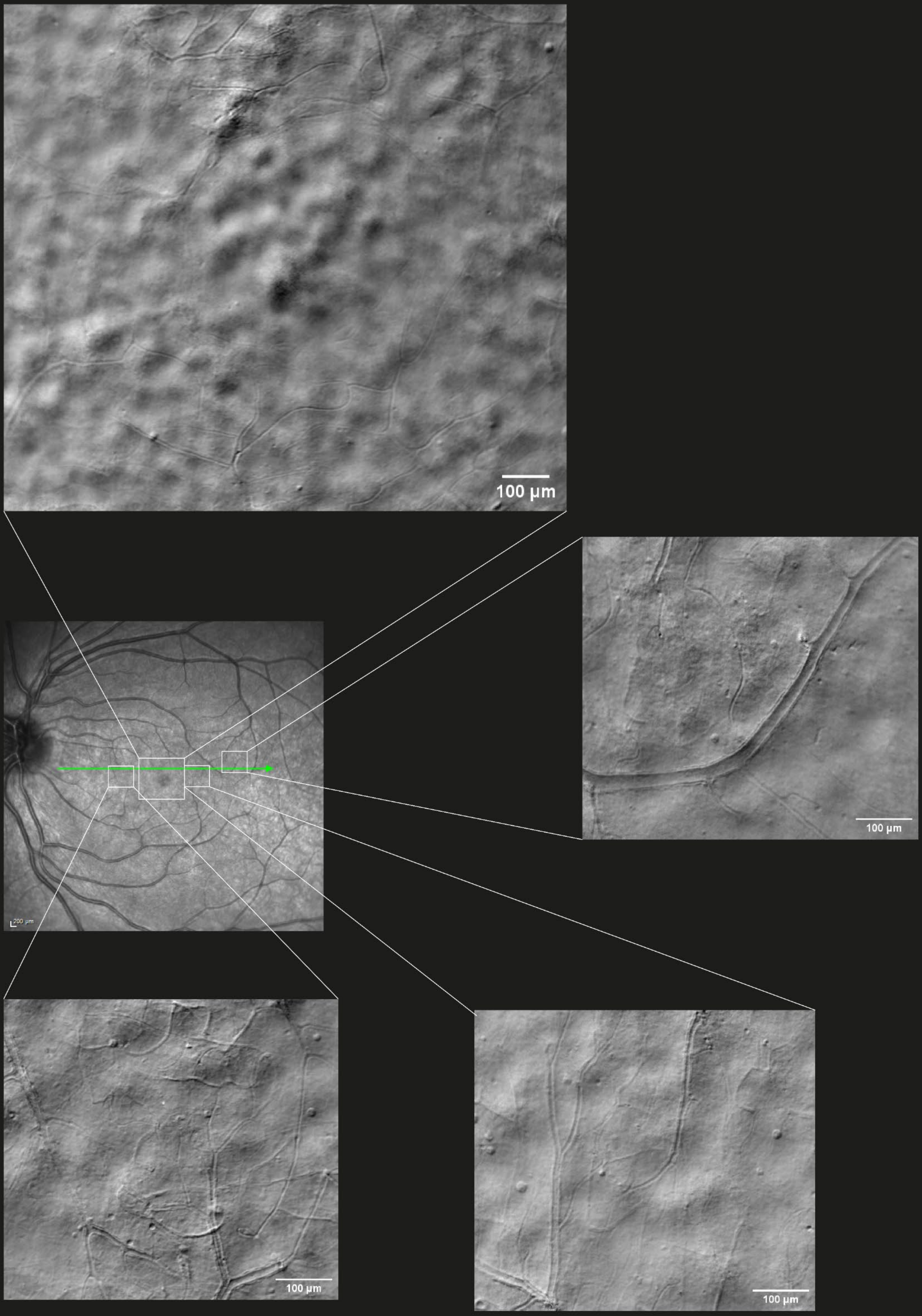


**Supplementary Figure 2.** Off-axis AOSLO images at retinal eccentricities beyond the study 4° temporal and nasal imaging protocol in an ON-RMS patient, revealing immune cells in additional retinal regions.

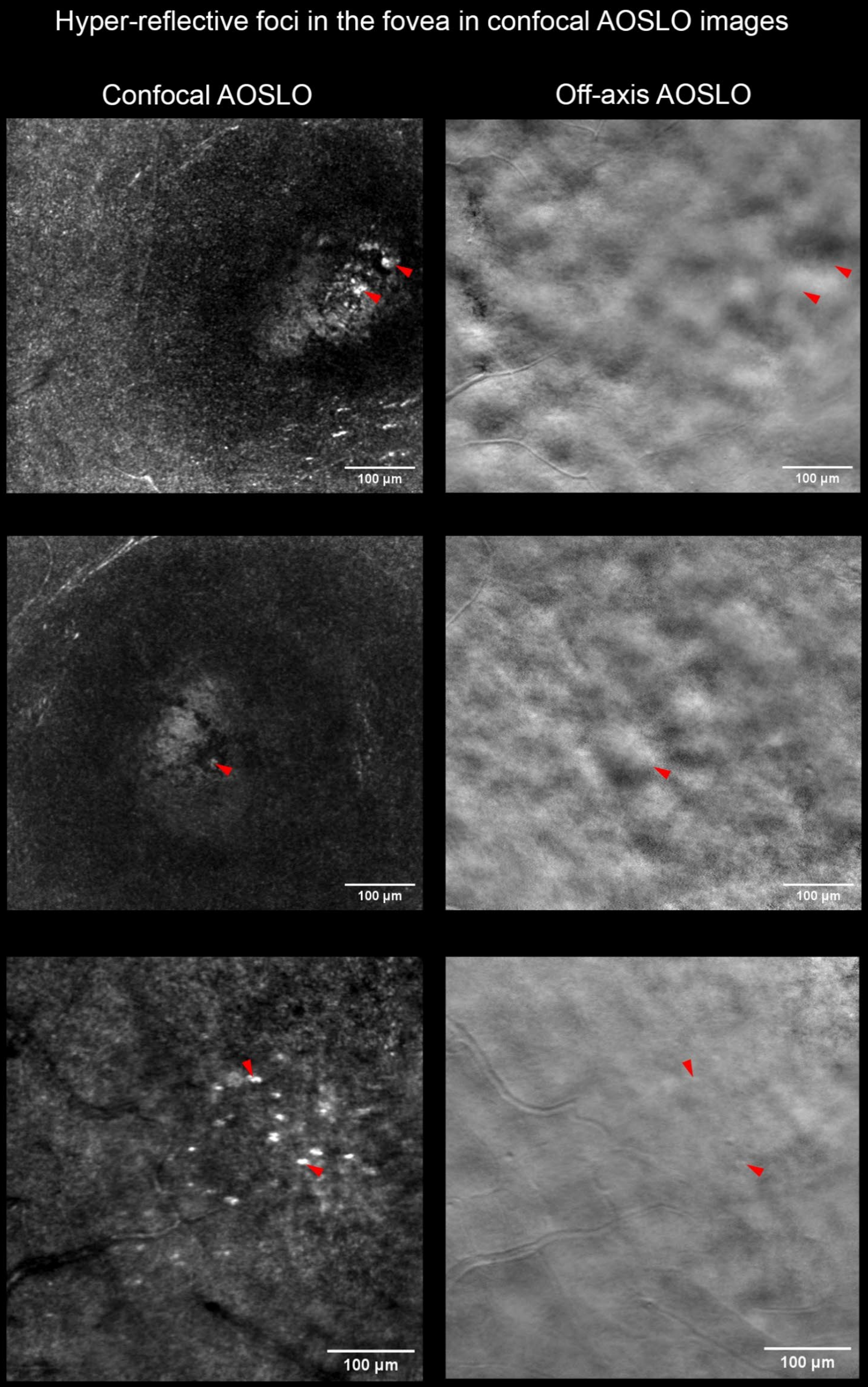


**Supplementary Figure 3.** (Left) Confocal AOSLO images of the fovea from three multiple sclerosis patients (one ON-RMS, two NON-RMS) showing hyperreflective dots previously described by Hargrave et al. (Hargrave A et al., Invest. Ophthalmol. Vis. Sci., 2020; 61(7):5101-5101). These structures do not produce a phase contrast signal in off-axis AOSLO images, indicating they are distinct from the cells described in this study.

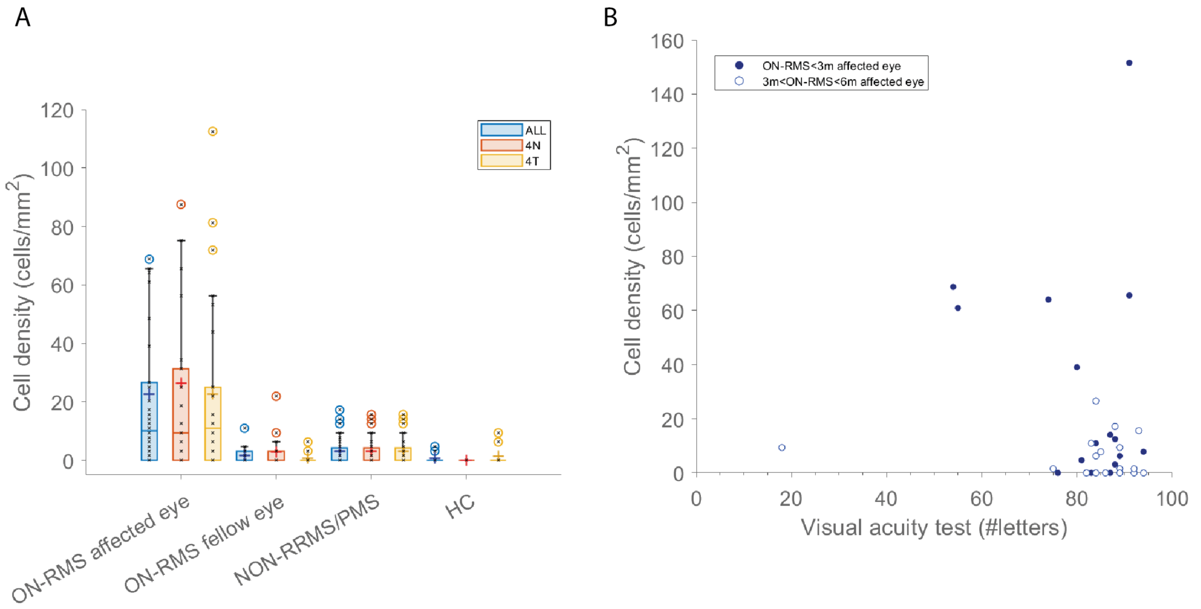


**Supplementary Figure 4.** (A) Influence of nasal vs. temporal retinal location on cell density at baseline (first AOSLO visit) across multiple sclerosis subtypes (ON-RMS: Relapsing MS with recent optic neuritis, NON-RMS: Relapsing MS without recent Optic Neuritis, PMS: Progressive MS) compared to Healthy Controls (HC). (B) Influence of visual acuity (low contrast) on cell density at baseline in patients from ON-RMS group.

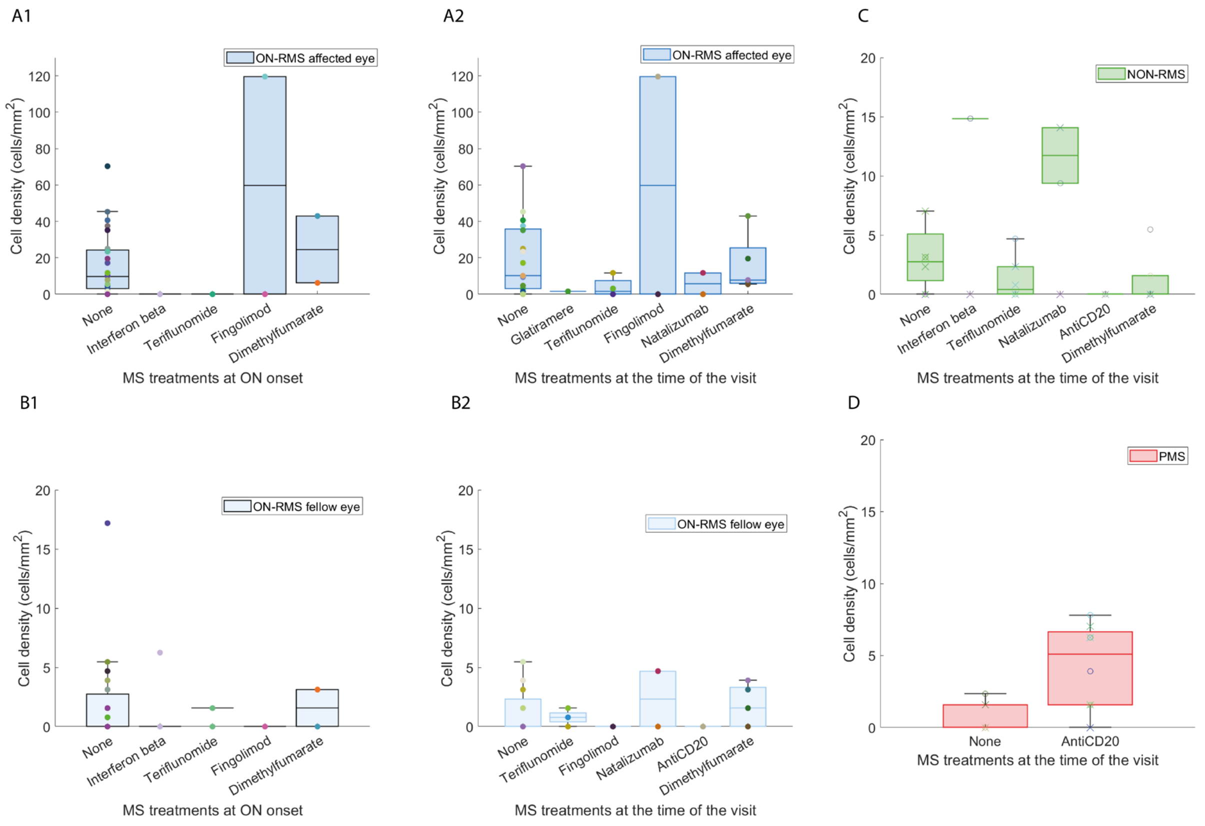


**Supplementary Figure 5**. Cell density at first AOSLO visit (baseline) plotted against disease-modifying treatments in multiple sclerosis (MS). (A1, B1) Cell density in the affected and fellow eyes of ON-RMS (relapsing-remitting MS with optic neuritis) patients, respectively, for different MS treatments at ON (optic neuritis) onset. (A2, B2, C, D) Cell density for different treatments at the time of the visit for ON-RMS affected and fellow eyes, as well as for NON-RMS (RMS without ON) and PMS (progressive MS) patients, respectively.

**Supplementary Table 1. Cells mean and median diameter for all MS groups and healthy controls**

| Group | Mean size (μm) | Median size (μm) | Standard Deviation (μm) |
|---|---|---|---|
| **ON-RMS affected eye** | 12.8 | 12.4 | 3.5 |
| **ON-RMS fellow eye** | 14.9 | 13.8 | 6 |
| **NON-RMS** | 11.1 | 11.1 | 2.7 |
| **PMS** | 10.3 | 10.7 | 1.7 |
| **HC** | 10.9 | 10.5 | 2.9 |

Acronyms: ON: Optic Neuritis; ON-RMS: Relapsing Remitting MS patients with recent optic neuritis; NON-RMS: Relapsing Remitting MS patients without recent optic neuritis; PMS: Progressive Multiple Sclerosis patients; HC: Healthy Subjects.
